# Predicting antibody kinetics and duration of protection against SARS-CoV-2 following vaccination from sparse serological data

**DOI:** 10.1101/2025.02.10.25322008

**Authors:** Julia Deichmann, Noam Barda, Michal Canetti, Yovel Peretz, Yael Ottolenghi, Yaniv Lustig, Gili Regev-Yochay, Marc Lipsitch

## Abstract

Vaccination against the severe acute respiratory syndrome coronavirus 2 (SARS-CoV-2) generates an antibody response that shows large inter-individual variability. This variability complicates the use of antibody levels as a correlate of protection and the evaluation of population- and individual-level infection risk without access to broad serological testing. Here, we applied a mathematical model of antibody kinetics to capture individual anti-SARS-CoV-2 IgG antibody trajectories and to identify factors driving the humoral immune response. Subsequently, we evaluated model predictions and inferred the corresponding duration of protection for new individuals based on a single antibody measurement, assuming sparse access to serological testing. We observe a reduced antibody response in older and in male individuals, and in individuals with autoimmune diseases, diabetes and immunosuppression, using data from a longitudinal cohort study conducted in healthcare workers at Sheba Medical Center, Israel, following primary vaccination with the BNT162b2 COVID-19 vaccine. Our results further suggest that model predictions of an individual’s antibody response to vaccination can be used to predict the duration of protection when serological data is limited, and could serve as a tool to estimate infection risk over time on both the population and individual level.

## Introduction

The introduction of vaccines against SARS-CoV-2 in late 2020 has played a pivotal role in combatting the global spread of COVID-19. They provide protection against severe disease and death [1, 2], and to a lower extent reduce the risk of infection and transmission [3, 4]. However, vaccine effectiveness and humoral immunity show a progressive decline with time after vaccination [5].

Vaccination against SARS-CoV-2 elicits a complex cellular and humoral immune response. The humoral immune response is characterized by a rise in antibody levels against the spike protein of the virus, and it was shown in population- and individual-level studies that they can serve as a correlate of protection against COVID-19 infection [6–8]. Antibodies initially rise and then fall rapidly in the first three months after vaccination. This is followed by a period of slow waning over several months [9], resulting in a biphasic pattern typical for antibody kinetics [10]. The waning of antibody levels is associated with an increase in infection risk over time, warranting repeated vaccination to maintain antibody levels and prevent the loss of protection.

However, antibody levels display high inter-individual variability. They are not only associated with time since vaccination, but also depend on other factors like age, sex and coexisting conditions [11, 12]. This large variation makes it challenging to predict antibody levels to evaluate population- and individual-level infection risk, diminishing their predictive value as a correlate of protection without broad and ongoing serological testing. It further limits their use in developing boosting strategies, especially when determining the optimal timing of vaccination in subpopulations with a reduced antibody response and shorter vaccine-induced duration of protection.

In this study, we aim to predict individual-level anti-SARS-CoV-2 antibody trajectories and the corresponding duration of protection following primary vaccination based on limited serological data. We rely on a mathematical model of antibody kinetics previously developed for malaria [13], which has later been applied in the context of SARS-CoV-2 [9, 14]. To quantify the effects of demographic characteristics and comorbidities on the antibody response, we calibrate the model using longitudinal IgG binding antibody data against the receptor-binding domain (RBD) of the SARS-CoV-2 spike protein. Data were collected in a cohort of healthcare workers at Sheba Medical Center, Israel, receiving two doses of the Bnt162b2 COVID-19 vaccine [15]. We then update the model to predict the antibody response of new individuals by providing a single serological measurement. We evaluate the effect of timing of serological sampling on prediction accuracy and finally determine the resulting duration of protection induced by vaccination.

## Results

### Data

Of the 4 868 healthcare workers in the study cohort, we included 2 609 with complete demographic information and at least two serological samples in our analysis, providing a total of 14 853 serum samples. Participant characteristics are summarized in Table 1. The cohort consists of 2 003 (77 %) female participants, with a median age of 47 years and BMI of 25 kg/m^2^. Participants exhibit few comorbidities. The most common comorbidity is hypertension in 290 participants (11 %), whereas immunosuppression only occurs in 36 participants (1 %). By design, no participants have a history of SARS-CoV-2 infection and all received two doses of the BNT162b2 COVID-19 vaccine.

**Table 1.** Characteristics of study participants overall and in the training and test groups.

|  | Total<br>(n=2609) | Training<br>(n=1957) | Test<br>(n=652) |
| --- | --- | --- | --- |
| Sex (female) | 2003 (76.8) | 1487 (76.0) | 516 (79.1) |
| Age (years) | 47 [38–57] | 47 [37–57] | 47 [38–57] |
| BMI (kg/m<sup>2</sup>) | 24.8 [22.1–27.8] | 24.8 [22.2–27.8] | 24.7 [22.1–27.9] |
| Weight (kg) | 69 [60–80] | 69 [60–80] | 68 [60–80] |
| **Comorbidities** |  |  |  |
| Hypertension | 290 (11.1) | 219 (11.2) | 71 (10.9) |
| Autoimmune disease | 186 (7.1) | 139 (7.1) | 47 (7.2) |
| Dyslipidemia | 177 (6.8) | 135 (6.9) | 42 (6.4) |
| Diabetes | 134 (5.1) | 111 (5.7) | 23 (3.5) |
| Lung disease | 93 (3.6) | 64 (3.3) | 29 (4.4) |
| Heart disease | 68 (2.6) | 53 (2.7) | 15 (2.3) |
| Immunosuppression | 36 (1.4) | 29 (1.5) | 7 (1.1) |

Reported as *n* (%) or median [IQR].

We split the data into training and test data sets, which consist of 75 % and 25 % of study participants, respectively (S1 Appendix, Sec. S1). Characteristics of the two subgroups remain similar (Table 1).

### Antibody trajectories

Anti-SARS-CoV-2 IgG antibody levels first increase and become detectable in week two after administration of the first vaccine dose (Fig. 1a, green). This is followed by a steep increase after administration of the second dose, with peak antibody levels sometimes exceeding the upper quantification limit of the antibody assay used here. Antibody levels then slowly wane over the remaining study period.

**Fig 1.**
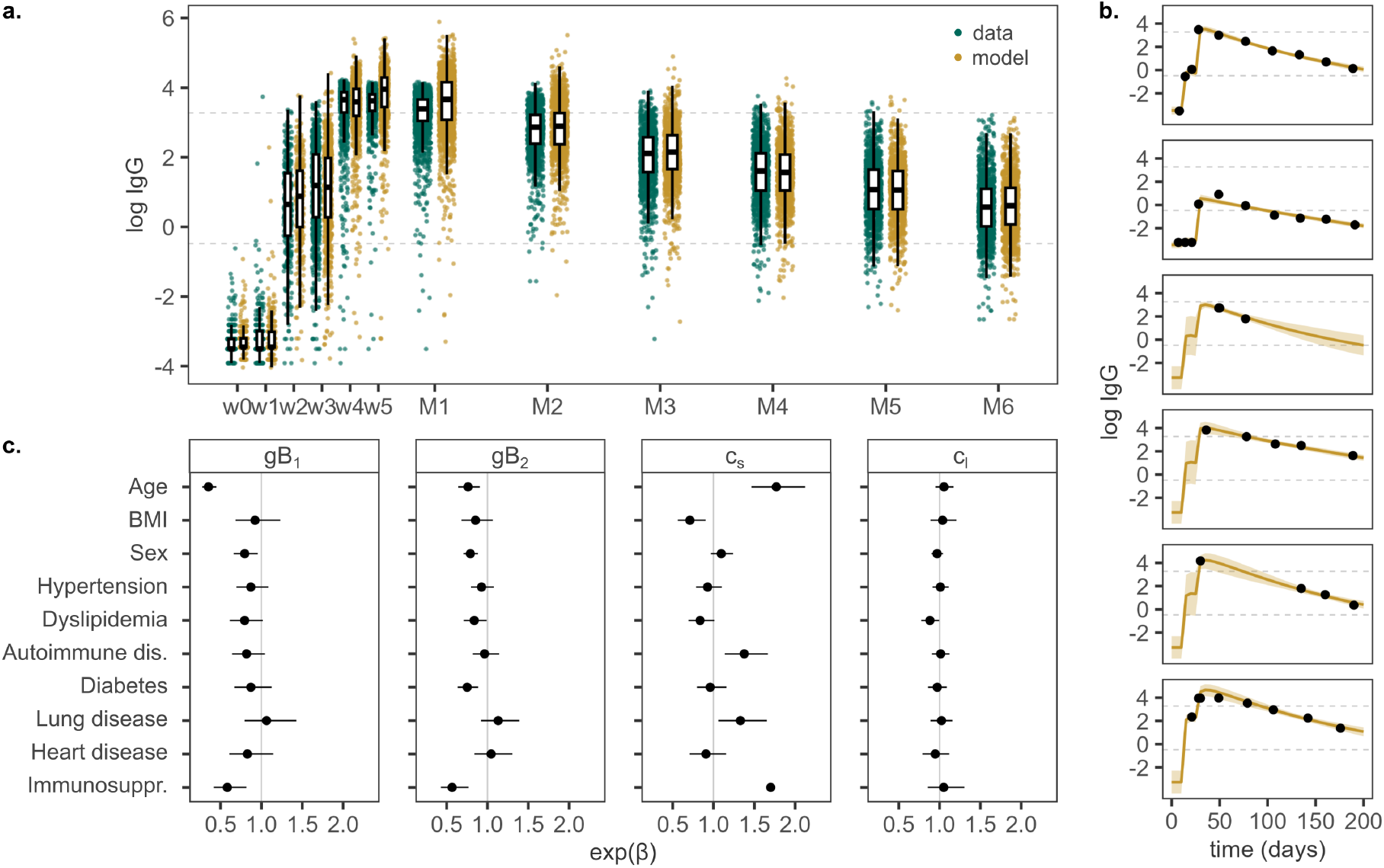
Model fit on the training data set. (a) Observed (green) and predicted (yellow) anti-SARS-CoV-2 IgG antibody levels following vaccination. Dose 1 is administered in week 0 (w0), dose 2 in week 3 (w3). Serum samples were taken weekly following dose 1 for five weeks (w1–w5), then monthly up to six months after dose 2 (M1–M6). The lower detection and upper quantification limits of the antibody assay are shown as dashed lines. (b) Observed and predicted antibody levels for six example individuals with different antibody response and sampling frequency. The solid line represents the geometric mean prediction, shaded areas indicate the 90 % posterior predictive interval. (c) Estimated effects of covariates on antibody boost (gB1 and gB2) and decay rate of short- and long-lived antibody-producing cells (cs and cl), shown as median and 95 % posterior interval estimates.

To describe immunological processes involved in the antibody response after vaccination and quantify differences between individuals, we fit a model of antibody kinetics to the individual-level antibody trajectories of the training data set. The model includes the antibody boost generated through vaccination by short- and long-lived antibody-producing cells (APCs), the decay of those cells and the waning of antibodies. We followed a Bayesian approach for model calibration, with individual parameter values originating from population-level parameter distributions. The inferred anti-SARS-CoV-2 IgG antibody levels capture the observed immune dynamics across the full training population and for individual study participants (Fig. 1a and b). Posterior distributions of the population-level parameters are normally distributed and a root mean squared log error (RMSLE) of 6.9 % (median, interquartile range [IQR]: 3.5–10.9 %) on the training data set indicates excellent agreement between data and model (S1 Appendix, Fig. S1). Peak antibody levels are reached 36 days (IQR: 34–37 days) after vaccination and the estimated half-life of the overall immune response is 69 days (IQR: 65–73 days), both showing little variation among individuals in the study population.

### Analysis of modeled antibody response

We further analyzed the modeled antibody trajectories and variability arising due to differences in demographics and comorbidities. In particular, we examined the amplitude of the immune response and the duration for which vaccination provides protective antibody levels (S1 Appendix, Fig. S2 and S3).

Antibody levels are lower for older individuals over the whole study period. This is also reflected in a decrease in maximum IgG levels with age, with a drop of 55 % (95 % confidence interval [CI]: 48–60 %) for participants over 60 years compared to participants younger than 30 years of age. While BMI does not affect the humoral immune response significantly, antibody levels are lower in male compared to female participants (ratio of means of peak IgG: 0.73 [95 % CI: 0.68–0.79]). Additionally, antibody levels are generally lower when coexisting conditions are present, in particular for autoimmune disease and diabetes (S1 Appendix, Fig. S3). There are 29 individuals in the training data set with immunosuppression, displaying the lowest antibody response to vaccination across the study population, with a decrease in peak antibody levels by 73 % (95 % CI: 65–79 %).

Vaccination induces a rise in antibody levels that provides protection against SARS-CoV-2 infection. However, the modeled duration of protection, defined as antibody levels over 500 BAU/mL [16] corresponding to a sample-to-cutoff ratio of 11.6 in our data [17], varies strongly between individuals (IQR: 75–115 days). In accordance with lower overall antibody levels, duration of protection decreases with age, where participants up to 30 years of age exhibit protective antibody levels for distinctively longer time periods than other age groups (S1 Appendix, Fig. S3). Similarly, the duration of protection is reduced in male compared to female individuals. Additionally, 78 out of 1957 (4 %) individuals remain unprotected over the full study period. This unprotected subpopulation consists mostly of older individuals and individuals with autoimmune diseases, diabetes and immunosuppression.

### Covariate effects on model parameters: antibody boost and APC decay

The variability in antibody response between individuals is captured in individual values for model parameters, which are associated with different processes involved in the humoral immune response. Here, we examined the effects of individual characteristics on the modeled antibody boost initiated by the first and second vaccine dose, represented by parameters *gB*_1_ and *gB*_2_, respectively, and the modeled decay of short-lived, *c*_*s*_, and long-lived antibody-producing cells, *c*_*l*_ (Fig. 1c).

The antibody boost decreases with age and is drastically reduced in older individuals following the first dose in particular, with a reduction by 52 % (95 % credible interval [CrI]: 43-59 %) from age 30 to age 60. In addition, we observe a lower antibody boost in male compared to female participants (factor *exp*(*β*), 0.80 [95 % CrI: 0.66–0.96] after dose one and 0.79 [95 % CrI: 0.71–0.88] after dose two) and in participants with dyslipidemia (0.80 [95 % CrI: 0.61–1.02] and 0.84 [95 % CrI: 0.71–0.99]) or immunosuppression (0.58 [95 % CrI: 0.42–0.82] and 0.57 [95 % CrI: 0.43–0.77]) after both vaccination events, and in people with diabetes after the second dose (0.75 [95 % CrI: 0.64–0.89]).

The decay rate of short-lived APCs increases with age (factor 1.49 [95 % CrI: 1.30–1.68] between age 30 and 60) and for individuals with autoimmune disease (1.38 [95 % CrI: 1.14–1.67]), lung disease (1.33 [95 % CrI: 1.06–1.65]) or immunosuppression (1.70 [95 % CrI: 1.27–2.45]). Consequently, the modeled half-life of short-lived APCs is shorter in older individuals and in the presence of these comorbidities. A higher BMI is associated with a reduction in decay rate (factor 0.92 [95 % CrI: 0.87–0.98] between a BMI of 22 kg/m^2^ and 28 kg/m^2^). In contrast, long-lived APCs remain unaffected by the examined covariates. Hence, the decay rate *c*_*l*_ and corresponding half-life are similar across the full study population.

### Prediction of antibody levels

For model validation, we predicted the antibody responses of individuals in the test data set. We provided one anti-SARS-CoV-2 IgG measurement per individual to adjust the model’s random effect estimates, assuming limited availability of serological testing. We evaluated both how well the model captures the full antibody trajectory as well as the accuracy of forward predictions from the time of testing. Moreover, we repeated this analysis with serological sampling being carried out at different times after vaccination to analyze the effect of timing on prediction accuracy.

We show predicted antibody trajectories for one example individual and residuals across the full test population in Fig. 2a and Fig. S4 (S1 Appendix), where IgG measurements are provided between month one (M1) and month six (M6) following vaccination. Overall, providing a single data point for model adjustment leads to an improved geometric mean prediction and refined posterior prediction interval compared to predictions made without model adjustment (S1 Appendix, Fig. S5). For serological testing of the individual participant during M1 and M2, antibody levels during waning are slightly overestimated (Fig. 2a). For the remaining sampling times, we observe good agreement between model predictions and measured IgG antibody levels, including narrower posterior prediction intervals. Similarly, residuals across the population are smaller for serological sampling performed during M3 or later compared to sampling performed during the first two months following vaccination, with the RMSLE between data and model predictions after administration of the second vaccine dose decreasing from 26.6 % (IQR: 13.7–49.0 %) for M1 to 18.0 % (IQR: 11.4–26.2 %) for M3 across the test population (Fig. 2a and S1 Appendix, Fig. S4). Afterwards, the RMSLE is stable for the remaining study period.

**Fig 2.**
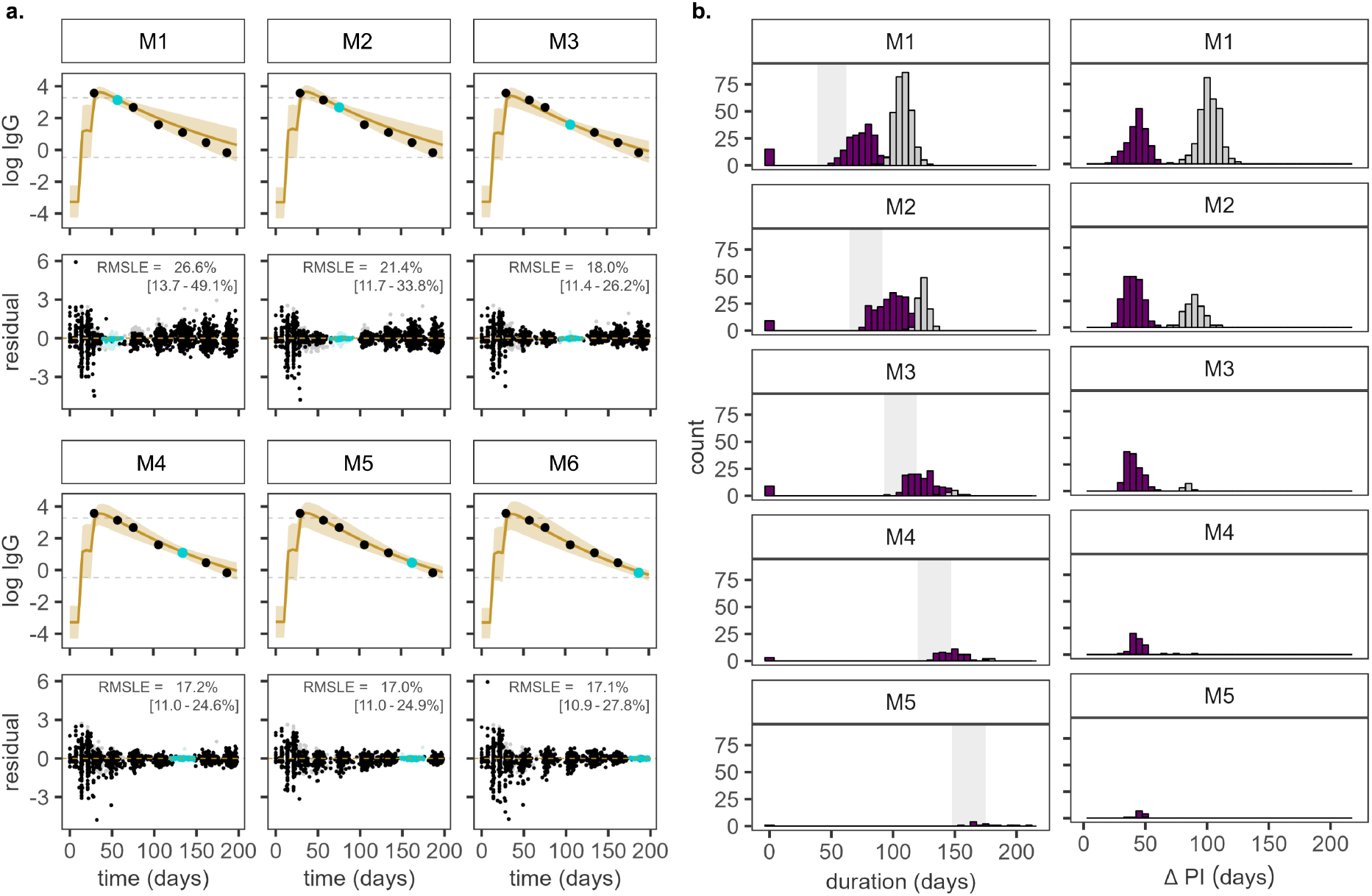
Prediction of antibody response on the test data set. (a) Predicted IgG antibody trajectories (yellow) for one example individual from the test data set. One measurement is provided between months one (M1) and six (M6) for model adjustment (light blue), the remaining data points (black) are unknown to the model. The solid line represents the geometric mean prediction, shaded areas indicate the 90 % posterior predictive interval. Residuals and the RMSLE for antibody levels after receipt of the second vaccine dose (median, IQR) are shown for the full test population, where lighter shades are used for measurements above the limit of quantification. (b) Predicted median duration of protection by sample time (left) and width of the 90 % prediction interval (PI, right) for measured antibody levels above the protective threshold. Individuals that never cross the protective threshold are represented with a duration of protection of 0. Purple and grey indicate that the provided measurement falls below or above the antibody assay’s upper limit of quantification, respectively.

Moreover, we quantified the accuracy of forward predictions from the time of serological testing in monthly increments using the RMSLE (Table 2). The antibody predictions one month ahead (diagonal, Table 2) are the most precise and comparable for all sampling times, with a median RMSLE between 14.5 % and 17.9 %. The RMSLE increases for predictions further into the future. Predictions one and two months ahead have a lower accuracy for sampling during M3, when the immune response transitions from short- to long-lived processes, compared to the surrounding months. Additionally, the variation in RMSLE among participants is higher for earlier sampling times and with increasing distance from the time of testing.

**Table 2.**
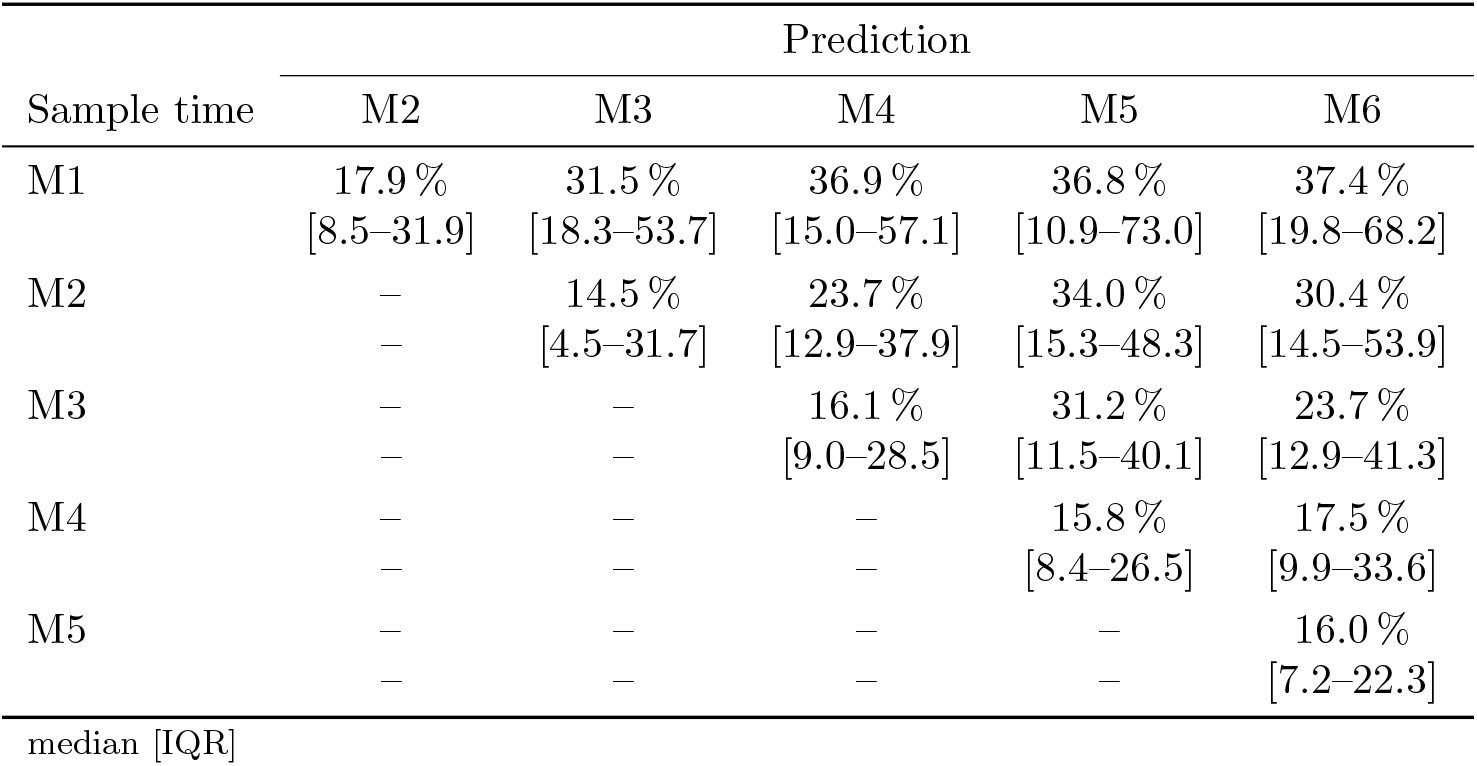
RMSLE between model predictions and test data for the months following serological sampling between month one (M1) and month five (M5).

### Prediction of duration of protection

Finally, we evaluated whether we can predict the time at which an individual’s antibody levels drop below the protective threshold, where we consider individuals that are protected at the time of testing. The distribution of the predicted duration of protection and corresponding uncertainty are displayed in Figure 2b for different sampling times between M1 and M6 after vaccination. We also identified individuals that never reach protective antibody levels.

The range of protective duration falls between 75 and 115 days (IQR) as determined on the training data set. For serological sampling in M1, we observe a similar distribution for the predictions on the test data set. For later sampling, only participants with an extended duration of protection are captured, since we assume that participants present antibody levels above the protective threshold at the time of testing. The width of the 90 % prediction interval remains stable around 40 to 45 days. Note, however, that the predicted duration of protection is longer for individuals with an IgG measurement above the antibody assay’s upper quantification limit, which is also associated with a higher uncertainty (Fig. 2b, grey). Other study participants are predicted to remain unprotected over the full study period with antibody levels never crossing the protective threshold.

We validated our predictions against the duration of protection inferred from the measured anti-SARS-CoV-2 IgG antibody levels (Table 3). In the majority of cases, we categorize individuals correctly into protected or unprotected. Furthermore, we are able to predict the time window when participants lose protection, where we achieve slightly higher accuracy when serological sampling is performed at later time points following vaccination.

**Table 3.** Validation of protection status and duration by sampling time between month one (M1) and month 5 (M5). We compare the number of unprotected (U) and protected (P) individuals between data and model prediction, and determine the proportion of protected whose duration of protection (t_prot_) is also predicted accurately. In this summary, individuals are excluded if the measured IgG antibody level for model calibration falls above the limit of quantification for the given month, and if the time window to extract the duration of protection from the data exceeds 45 days.

| Data | Prediction |  |  |  |  |  |  |  |  |  |  |  |  |  |  |
| --- | --- | --- | --- | --- | --- | --- | --- | --- | --- | --- | --- | --- | --- | --- | --- |
|  | M1 (n=145) |  |  | M2 (n=173) |  |  | M3 (n=104) |  |  | M4 (n=38) |  |  | M5 (n=8) |  |  |
|  | U | P | (t<sub>prot</sub>) | U | P | (t<sub>prot</sub>) | U | P | (t<sub>prot</sub>) | U | P | (t<sub>prot</sub>) | U | P | (t<sub>prot</sub>) |
| U | 13 | 3 |  | 6 | 0 |  | 6 | 0 |  | 2 | 0 |  | 1 | 0 |  |
| P | 2 | 127 | (71%) | 2 | 165 | (82%) | 1 | 97 | (64%) | 1 | 35 | (86%) | 0 | 7 | (100%) |

## Discussion

IgG binding antibody levels against SARS-CoV-2 have been used as a correlate of protection for COVID-19 infection. However, this relies on access to broad serological testing, since large inter-individual variability in the humoral immune response to vaccination or infection makes it difficult to predict antibody levels and evaluate the population- and individual-level infection risk. In this study, we used a model of antibody kinetics to capture the antibody response to vaccination and identify factors that lead to differences between individuals. We then evaluated the prediction accuracy of the model and its capability to predict the time until a threshold IgG level is reached.

We followed a Bayesian approach to infer model parameters using antibody data collected in healthcare workers after primary vaccination. We quantified the effects of individual characteristics on parameters describing the vaccine-induced antibody boost and the decay rate of short- and long-lived antibody-producing cells, which govern the amplitude and duration of the antibody response. The model accurately describes individual antibody trajectories in the training data set. While the half-life of the overall response is similar across the full study population, we observe differences in amplitude and the duration of protection.

Many of the factors associated here with a lower vaccine-induced antibody boost and shorter half-life of APCs, yielding lower antibody levels, are known to impair the humoral immune response to vaccination and increase the risk of severe acute COVID-19 [11, 12, 18]. Older age is associated with a lower antibody response [19, 20]. In our study, this is captured in a reduction in antibody boost, especially following the first vaccine dose. This finding is in agreement with a decreased likelihood of seropositivity in individuals 60 years of age and older before receiving the second dose of the BNT162b2 COVID-19 vaccine [21]. In combination with a shorter modeled half-life of short-lived APCs, this leads to a decrease in antibody levels with age over the whole study period, and consequently a shorter duration of protection. Similar to other studies, we observe a stronger antibody response in female compared to male study participants [22], driven by a higher antibody boost after both vaccine doses. The modeled half-life of short-lived APCs increases with BMI. However, we do not detect significant differences in amplitude or duration of the resulting antibody response for different BMI categories. Although obesity has been linked to a higher rate of hospitalization and death from COVID-19 [23], IgG antibody levels were shown to be similar between individuals with severe obesity and a normal BMI. Instead, obesity was associated with a lower neutralizing capacity of these antibodies [24]. A lower antibody response is further associated with autoimmune disease and diabetes [11]. For both comorbidities, the duration of protection is reduced and the proportion of individuals that never reach protective antibody levels following the primary vaccination series is elevated. Finally, our results capture the impaired antibody response due to immunosuppression [11, 25]. A significant reduction in antibody boosting following vaccination and the half-life of short-lived APCs leads to a drastic reduction in maximum IgG levels and a high proportion of unprotected individuals.

Next, we applied the model to participants in the test data set that was not used for model inference.

It accurately predicts individual IgG antibody trajectories after updating the model based on a single serological sample to mimic limited access to serology. The highest prediction accuracy for the full antibody trajectory is achieved when testing is performed during antibody waning in month three following vaccination or later. Forward predictions perform well for all sampling times and the variability in prediction accuracy across the study population decreases for later sampling. Consequently, we are also able to predict the duration of protection of individuals who display protective antibody levels at the time of testing, suggesting that serological testing during month two following vaccination is most beneficial, when the majority of individuals display protective antibody levels and prediction accuracy is high. In contrast to previous studies, where antibody levels are projected forward following a longitudinal serology period used for model calibration [14, 26], these individual-level predictions rely on minimal data. While they give insight into an individual’s infection risk, this approach can also be leveraged on the population level to assess the duration of protection against infection across the full or different subpopulations and could thus be used to inform public health decisions on booster vaccination.

Higher antibody levels are generally associated with a lower risk of infection with COVID-19 [27]. In this study, we used an antibody threshold of 500 BAU/mL to define protection. However, the protective antibody threshold varies for different variants of concern [28]. With the emergence of immune-escaping variants, as we have seen during the spread of Omicron, higher antibody levels may be required to protect against infection. To adapt our analysis and evaluate the duration of protection against different variants, variant-specific antibody thresholds could be utilized. Alternatively, we could extend our model to link antibody kinetics and neutralizing capacity, for example by assuming a dose- and variant-dependent proportional relationship between antibody levels and neutralization [26]. In this case, the duration of protection could be inferred directly from the predicted neutralizing capacity based on a common neutralization threshold.

There are several limitations to this study. We used data from the Sheba healthcare worker cohort study, which consists of adults between 18 and 82 years of age. While a wide age range is covered and common comorbidities are represented, the proportion of older individuals and individuals with comorbidities in the cohort is lower compared to the general population. In addition, study participants all received the vaccine at the same time, with similar timing between doses and no prior COVID-19 infection, warranting care when transferring our results to a general, more heterogeneous population.

After administration of the second vaccine dose, measurements partially exceeded the antibody assay’s dynamic range. The model allowed us to infer peak antibody levels. However, censoring led to increased uncertainty in model parameters and antibody predictions.

Furthermore, this study is limited to the humoral immune response following the primary vaccination series against SARS-CoV-2. To predict antibody kinetics and duration of protection at present, boosting through both repeated vaccination and infection would need to be captured by the model. It was shown that the timing and number of booster vaccinations affect antibody kinetics and infection risk [17, 29], and that hybrid immunity from vaccination and previous infection yields a higher antibody boost and provides improved protection against symptomatic infection [30]. Additionally, our analysis is limited to binding antibodies. However, it was shown that the correlation between binding and neutralizing antibodies has decreased with the changing variants [31], impeding the use of binding IgG antibody levels as a correlate of protection. Consequently, timing, number and order of boosting events would need to be considered as additional model factors modulating the anti-SARS-CoV-2 antibody response, including model calibration on both binding and neutralizing antibody data.

Binding antibody levels play a critical role in monitoring the infection risk for SARS-CoV-2. However, this is challenged by the waning of the antibody response and high variability between individuals that may be difficult to capture without continuous serosurveillance. In this study, we demonstrated how model predictions of an individual’s antibody response to vaccination could be used to predict the duration of protection when serological data is limited. While we focused on primary vaccination, future work taking into account repeated vaccination, infection history and neutralization may benefit from the approach presented here. The approach further provides a platform for future pandemics, and could be applied as a tool to estimate population- and individual-level infection risk over time and support decision-making for vaccination strategies.

## Methods

### Data

We used data from a prospective longitudinal cohort study conducted in 4 868 healthcare workers at Sheba Medical Center, Israel [15]. Serum samples were collected before participants received the first dose of the BNT162b2 COVID-19 vaccine in December 2020. Administration of the first vaccine dose was followed by weekly serological testing for five weeks. The second dose was administered during week three in January 2021. Afterwards, serum samples were collected monthly. To be included in this study, healthcare workers had to have a negative anti-SARS-CoV-2 IgG assay before receiving the first dose. A positive SARS-CoV-2 PCR test or a positive anti-SARS-CoV-2 IgG serology test before vaccination led to exclusion from the study. Samples of individuals with COVID-19 infection after vaccination were included up to the positive test and post-infection measurements were excluded.

Participating healthcare workers completed a questionnaire to collect information on demographics and comorbidities. We removed all participants with missing information and less than two serological samples from our analysis. We further split the study population into a training and test data set (S1 Appendix, Sec. S1).

### Immunoassay

IgG levels against the receptor-binding domain (RBD) of the SARS-CoV-2 spike protein were measured using the Access SARS-CoV-2 RBD IgG assay (Beckman-Coulter, CA, U.S.A.). Seropositivity was defined by an IgG sample-to-cutoff (s/co) ratio greater than 0.62 [32]. In addition, we determined the upper limit of quantification (26.4 s/co) from serum samples collected in the month following administration of the second vaccine dose (days 25–38). During this period, the majority of measurements fall outside the assay’s dynamic range, and we used the median minus one standard deviation to define the quantification limit.

### Model of antibody kinetics

We describe antibody kinetics A(t) following vaccination using a mathematical model that captures the generation of antibodies by APCs and the subsequent waning of the antibody response [13]. The model is defined as

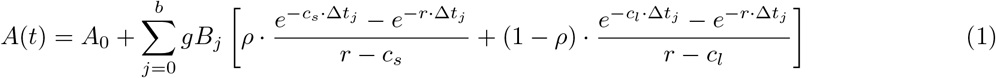

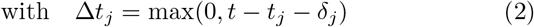

and initial antibody concentration *A*_0_. We allow for multiple boosting events j at times *t*_*j*_ due to repeated vaccination [14]. Each event is characterized by the antibody boost *gB*, representing the overall antibody production by APCs, and the delay in the humoral response *δ* after exposure. A proportion *ρ* of APCs are short-lived with decay rate *c*_*s*_, while a proportion (1 − *ρ*) are long-lived with decay rate *c*_*l*_, generating an antibody response with a bi-phasic waning phase. Finally, antibodies decay at rate *r*.

We estimated model parameters on longitudinal IgG measurements from study participants in the training data set. We defined parameters using mixed-effects models to account for variation in antibody response between individuals, taking into consideration age, sex, BMI and coexisting conditions. The model was then fitted in a Bayesian hierarchical framework. More details on the statistical model and inference are provided in the Supplementary Material (S1 Appendix, Sec. S2).

### Statistical analysis

We first evaluated the individual-level anti-SARS-CoV-2 antibody response based on the inferred antibody trajectories. We quantified the maximum IgG antibody level and examined the effects of demographic covariates and comorbidities in a multivariate linear regression. Similarly, we extracted and analyzed the duration of protection against infection provided by the initial vaccination series, where we defined the duration of protection as the time it takes for IgG levels to fall below a threshold previously determined to be protective against the delta variant [16].

Next, we applied the model to study participants in the test data set. To evaluate its prediction accuracy in the case of limited availability of serological data, we provided one measurement per individual to update the model and predict IgG antibody trajectories (S1 Appendix, Sec. S3). We then evaluated the overall accuracy of the model prediction and the accuracy of forward predictions in monthly increments using the root mean squared log error (RMSLE). Furthermore, we predicted the duration of protection for each individual that showed protective antibody levels at the time of serological testing. We assessed the uncertainty associated with these predictions and validated them against the duration of protection inferred from the available longitudinal IgG measurements (S1 Appendix, Sec. S4).

## Data Availability

--

## Supporting information

### S1 Appendix

Supplementary Methods and Figures

## Acknowledgments

The authors thank David Helekal for feedback on the model implementation and Rahul Subramanian for valuable discussions.

