## Supplementary material for "Predicting antibody kinetics and duration of protection against SARS-CoV-2 following vaccination from sparse serological data": S1_Appendix

### Supplementary Methods

#### S1 Training and test data

We used serum samples from 75 % of the study population as a training data set for model inference and used samples from the remaining 25 % as a test data set for the evaluation of model predictions. In order to have data from participants with underlying conditions present in both data sets, we divided the population into groups with and without comorbidities, and randomly assigned individuals to training and test sets separately for both groups.

The characteristics of the full study population as well as summaries for the training and test sets are provided in Table 1 in the main text.

#### S2 Statistical model and inference

##### S2.1 Observation model

We assume that the observed antibody levels $y_{ij}$ of individual $i$ at time $j$ are log-normally distributed with $y_{ij} \sim LogN(A_{ij}, \sigma)$, where $A_{ij}$ represent the modeled antibody levels. The measurement error $\sigma$ is estimated with the remaining parameters during model inference and prior $\sigma \sim N_+(0, 0.1)$.

Antibody levels are imputed for censored observations above the limit of quantification, $y_{max}$. Similar to uncensored observations, we assume that they follow a log-normal distribution around the model predicted values. This implies that $\hat{y}_{ij} \sim LogN(A_{ij}, \sigma)$, with the imputed value $\hat{y}_{ij}$ and $\hat{y}_{ij} > y_{max}$.

##### S2.2 Individual-level parameters

The antibody kinetics model comprises nine immunological parameters that need to be estimated. We use mixed effects models to infer individual-level parameter values and capture the variability between study participants in the corresponding immunological processes, while estimating the distribution of these parameters across the entire population. For the antibody boost following the first and second dose, $gB_1$ and $gB_2$, respectively, and the decay rates of short-lived, $c_s$, and long-lived antibody-producing cells (APCs), $c_l$, these mixed effects models take on the general form:

$$\begin{aligned} log(p_i) &= log(p_{pop}) + X_i^T \cdot \beta_p + \epsilon_{p,i} \qquad && \epsilon_{p,i} \sim N(0, \sigma_p) \\ &= log(p_{pop}) + X_i^T \cdot \beta_p + u_{p,i} \cdot \sigma_p && u_{p,i} \sim N(0, 1), \end{aligned} \tag{1}$$

where $p_i$ represents parameter $gB_1$, $gB_2$, $c_s$ or $c_l$ of individual $i$. The intercept term $log(p_{pop})$ describes the average parameter value $p_{pop}$ across the whole population. The fixed effects term $X_i^T \cdot \beta_p$ captures deviations from the population average that can be explained by individual characteristics, summarized in a $k$-dimensional covariate vector $X_i$, with slope vector $\beta_p$ that is common between all individuals. Finally, the remaining inter-individual variability is captured in the random effects term $u_{p,i} \cdot \sigma_p$, which consists of the individual deviation $u_{p,i}$ in standard deviations $\sigma_p$.

Age, BMI and sex (female=0, male=1) are added as fixed effects to the covariate matrix $X$. In addition, we include the comorbodities listed in Table 1 in the main text. We re-scale and take the logarithm of the continuous covariates age and BMI, such that the population parameters are defined for a typical female of 43 years with a BMI of $25\,\mathrm{kg/m^2}$ and no comorbidities.

We assume that there are no systematic differences in initial antibody levels between individuals, since all participants have a negative anti-SARS-CoV-2 IgG assay before entering the study. We therefore removed fixed effects for parameter $A_0$. Similarly, we assume that antibodies have a decay rate $r$ that is consistent across the population. Consequently, differences in the waning of antibody levels will be captured in the decay rates of APCs. Since $\rho$ describes a proportion and is bound between 0 and 1, we define $\rho_i = logit^{-1}(\rho_{pop} + u_{\rho,i} \cdot \sigma_\rho)$.

We fix the delay in antibody response following vaccination based on the time we first observe an increase in antibody levels in the data. This results in $\delta_1 = 10$ days for dose one and $\delta_2 = 4$ days for dose two. While we allow the antibody boost $gB_j$ and delay $\delta_j$ to be different for each vaccine dose $j$, we assume that $\rho$ and all decay rates remain the same, since potential differences cannot be inferred due to the proximity of the two vaccination events relative to processes with longer time scales involved in the humoral response.

#### S2.3 Priors

We defined the following priors for population- and individual-level parameters:

$$
\begin{aligned}
log(A_{0,pop}) &\sim N(log(0.05), 0.5)\\
log(gB_{1,pop}) &\sim N(log(0.8), 0.2)\\
log(gB_{2,pop}) &\sim N(log(10), 0.2)\\
\rho_{pop} &\sim N(3, 0.1)\\
log(r) &\sim N(log(0.033), 0.1)\\
log(c_{s,pop}) &\sim N(log(0.14), 0.1)\\
log(c_{l,pop}) &\sim N(log(0.002), 0.5)\\
\beta_{[gB1/gB2/cs/cl]} &\sim N(0, 0.2)\\
u_{[A0/\rho/gB1/gB2/cs/cl]} &\sim N(0, 1)\\
\sigma_{[A0/\rho/gB1/gB2/cs/cl]} &\sim N_{+}(0, 0.3)
\end{aligned}
\tag{2}
$$

Parameters $A_0$, $gB_1$ and $gB_2$ are related to the amplitude of the antibody response and are therefore assay-specific. We used a prior on $\rho$ that corresponds to a high proportion of short-lived APCs [1,2]. We set priors for rate parameters based on antibody and APC half-lives. IgG antibodies have a half-life of $t_{1/2} = 21$ days [3], which relates to their decay rate via $r = log(2)/t_{1/2}$. Short-lived APCs have a half-life of only a few days [4]. Here, we set the prior according to $t_{1/2} = 5$ days. The half-life of long-lived APCs, on the other hand, remains uncertain and we chose a wider prior for the corresponding rate parameter.

#### S2.4 Inference

We sampled from the posterior distributions using the No-U-Turn sampler (NUTS) [5], a Markov Chain Monte Carlo (MCMC) algorithm implemented in Stan [6]. We used 1000 warm-up iterations, 2000 sampling iterations and four chains. No divergences were detected and estimated parameters had $\hat{R} \leq 1.03$, indicating that chains have mixed well. Processing of results was performed in R with the CmdStanR package.

### S3 Prediction of antibody levels on test data

Next, we aimed to predict the anti-SARS-CoV-2 antibody response of a new individual that was not part of the training data set based on limited serological data. We considered study participants from the test data set and provided one IgG measurement per individual.

To predict antibody levels, we first updated the model given the new data. The model parameters consist of a population average, fixed and random effects. The first two components are described by population-level parameters. These were determined on the training data set and we can re-use the posterior draws for participants in the test data set. The random effects, however, contain an unknown deviation $u_{p,i}$ for parameter $p$ and individual $i$. We sampled the deviation $u_{p,i}$ from a standard normal distribution (see Eq. 1) and simulated the corresponding IgG antibody trajectories. We then selected realizations that generate predictions such that the available observation falls into the 90 % posterior prediction interval of the observation model.

Model predictions were evaluated using the root mean squared log error (RMSLE) based on all observations available for an individual. In addition to the general prediction accuracy, we are also interested in the accuracy of forward predictions over varying time horizons and their dependence on the timing of serological testing after vaccination. Since testing was performed monthly after administration of the second vaccine dose, we provided a sample from month one (M1: days 39–63 following the first dose [7]), two (M2: days 64–91), three (M3: days 92–119), four (M4: days 120–147), five (M5: days 148–175) or six (M6: days 176–203) for model adjustment and analyzed the RMSLE of the predicted antibody response separately for the subsequent months.

### S4 Duration of protection

Moreover, we examined the ability of the model to predict the duration of protection provided by the vaccine. We defined an anti-SARS-CoV-2 IgG antibody level of 500 BAU/mL, corresponding to a sample-to-cutoff ratio of 11.6 in our data [8], as the threshold for protection. This threshold was previously determined for protection against the delta variant using this data set [9]. We evaluated the time at which simulated antibody levels fall below the threshold and report the median duration of protection for each individual, including the width of the 90 % posterior prediction interval to quantify uncertainty.

We considered individuals that have a median duration of protection larger than 0 as protected, meaning that at least 50 % of sampled antibody trajectories cross the protective threshold. The remaining individuals are considered to remain unprotected following vaccination.

#### S4.1 Validation

For participants in the test data set, we compared our predictions against the observed duration of protection. Initially, we validated our categorization into protected and unprotected. Afterwards, we validated the predicted duration for protected individuals. To do so, we evaluated whether our prediction falls into the time interval between the last data point above and the first data point below the protective threshold during the waning phase of the antibody response. Note that we included participants only if this time interval does not exceed 45 days to avoid overestimating the prediction accuracy of our model due to sparse serological sampling.

### Supplementary Figures

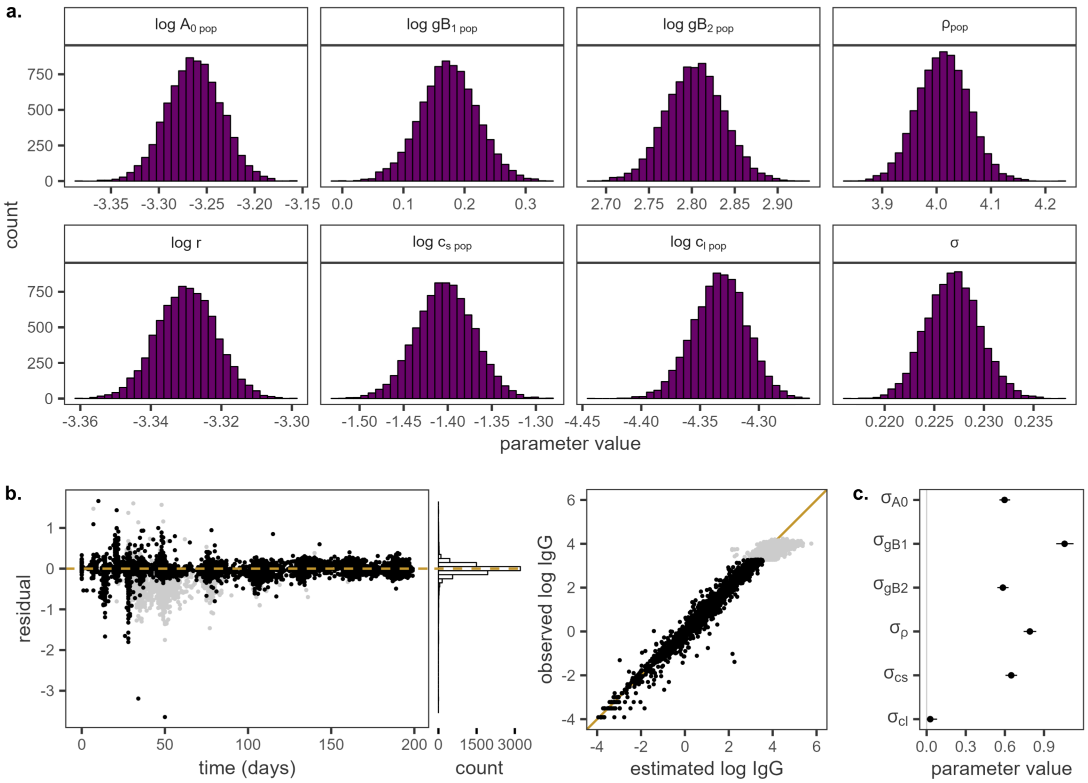


**Fig S1. Parameter estimates and residuals.** (a) Posterior distributions of the estimated population-level parameters. (b) Residuals over time and comparison of observed and estimated IgG levels. Grey points indicate measurements above the antibody assay's limit of quantification. (c) 95 % interval estimates of the random effect parameters.

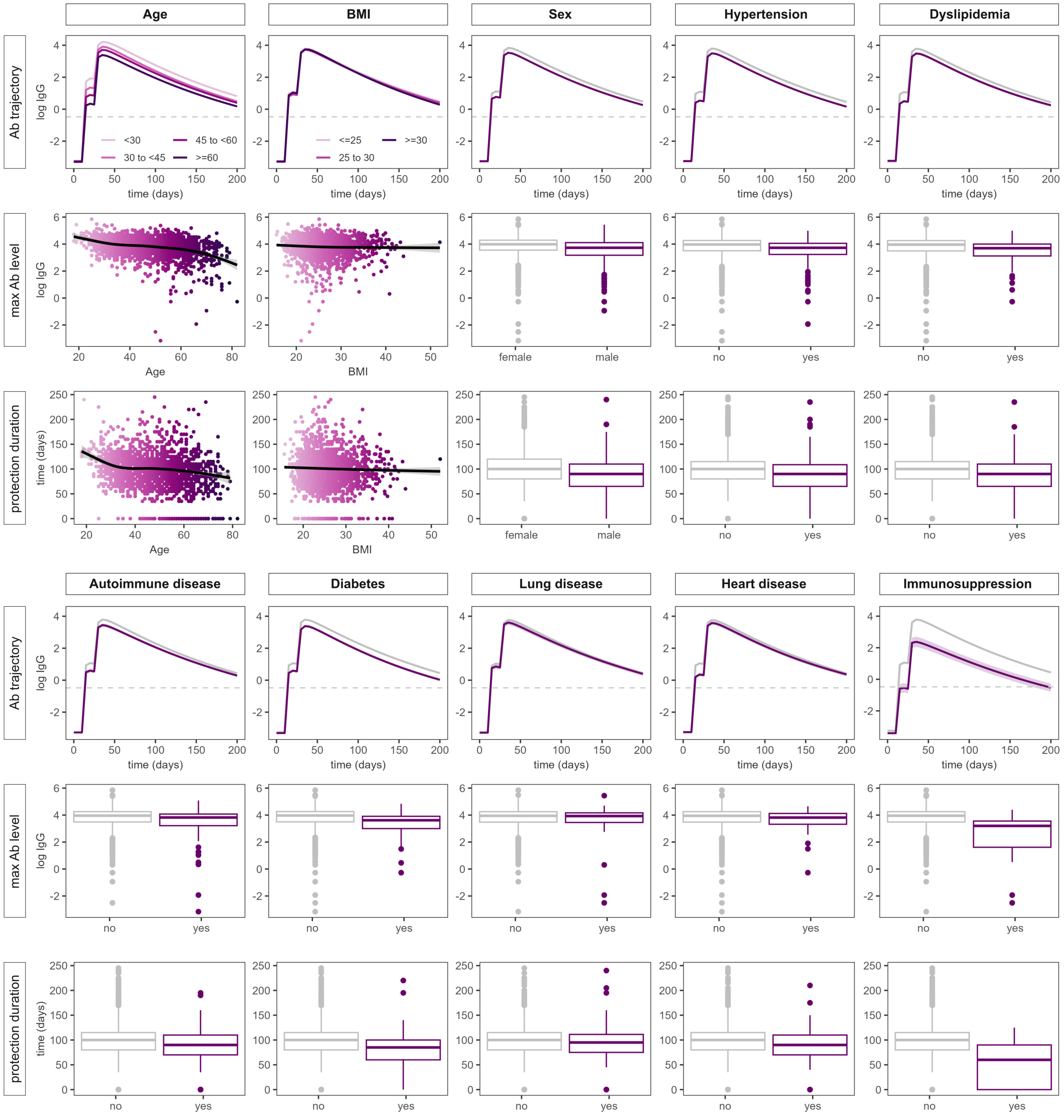


**Fig S2. Evaluation of antibody response.** Predicted antibody trajectories, maximum IgG antibody level and duration of protection stratified by the covariates considered in the antibody kinetics model.

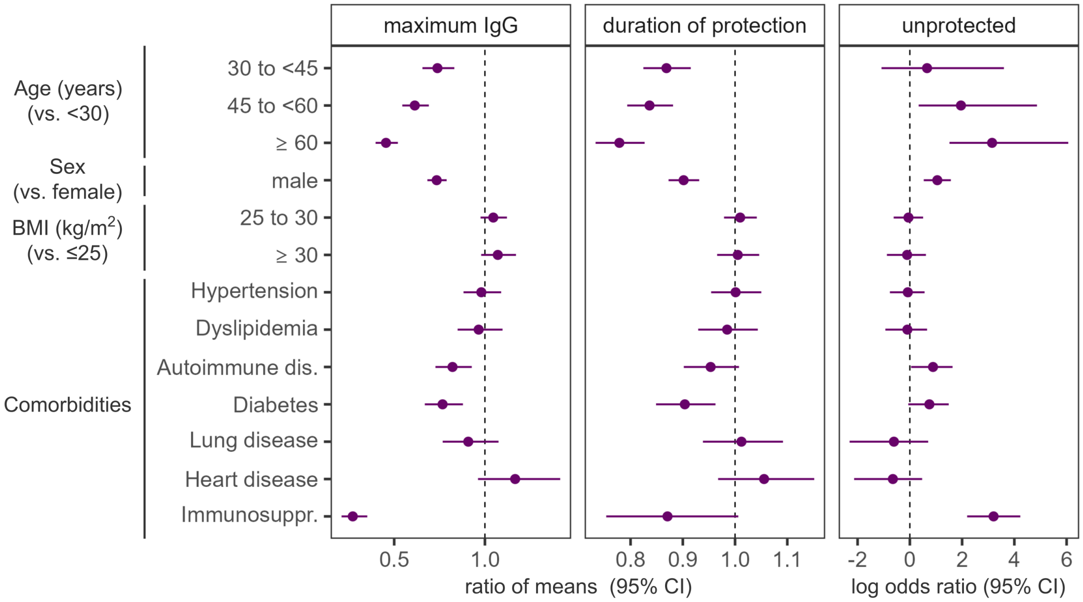


**Fig S3. Analysis of maximum IgG levels and duration of protection** from modeled antibody trajectories using multivariate linear mixed models, and log odds ratio of remaining unprotected following vaccination. Linear regression is performed on log antibody levels and log duration. We show exponentiated $\beta$ coefficients, representing the ratio of means between the examined factor and the reference level.

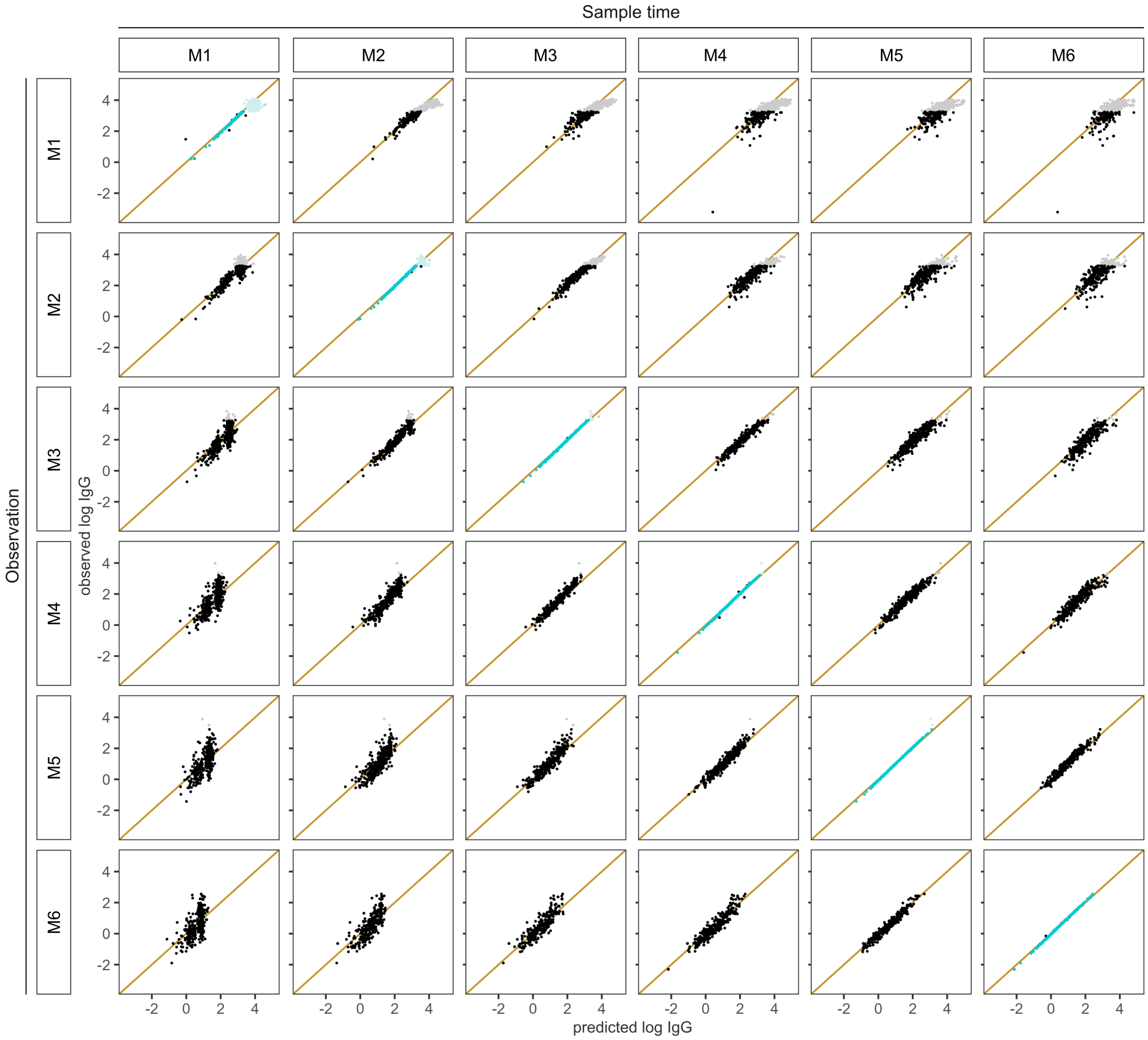


**Fig S4. Observed vs predicted antibody levels** by sample time between M1 and M6 (columns) used to update the random effect parameters of the model for individuals in the test data set, and time of observation (rows). The provided data points are shown in blue, the remaining data points (black) are unknown to the model. Lighter shades indicate observations above the limit of quantification.

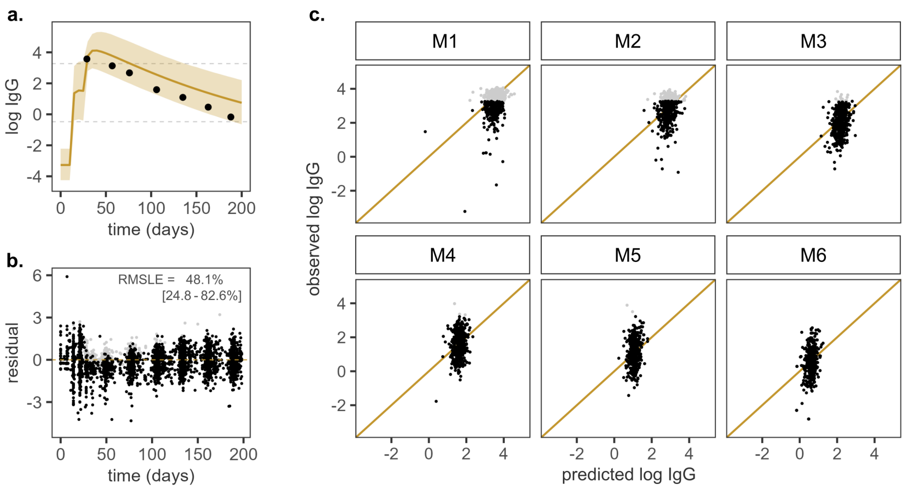


**Fig S5. Prediction of antibody levels without model adjustment.** (a) Predicted IgG antibody trajectory (yellow) for one example individual from the test data set without model adjustment. The solid line represents the geometric mean prediction, the shaded area indicates the 90 % posterior predictive interval. (b) Residuals and the RMSLE for antibody levels after receipt of the second vaccine dose (median, IQR) are shown for the full test population. (c) Observed vs predicted antibody levels by time of observation between M1 and M6, where geometric mean predictions display lower inter-individual variability than the observed data. Lighter shades are used for measurements above the limit of quantification.
